# Annual HIV incidence among gay, bisexual and other men who have sex with men in England, 2014 to 2023: A prospective cohort analysis using national surveillance data

**DOI:** 10.1101/2025.07.02.25330732

**Authors:** Natasha Ratna, Catriona Harrison, Tobi Kolawole, Eleanor Bell, Dana Ogaz, Alison Brown, John Saunders, Tamara Djuretic, Hamish Mohammed

## Abstract

Following the fall in new HIV diagnoses in gay, bisexual and other men who have sex with men (GBMSM) since 2015, the English government published an HIV Action Plan committing to end HIV transmission by 2030. Underlying HIV transmission, migration and access to testing, influence diagnosis trends and may not reflect incidence as people could be living with undiagnosed HIV for many years. We derived HIV incidence by clinical risk markers and PrEP use.

Using GUMCAD STI surveillance data between 2014/15 and 2022/23, we calculated yearly HIV incidence among HIV-negative GBMSM attending sexual health services (SHS) in England with at least 2 HIV tests within 365 days (‘repeat testers’). Annual incidence was stratified by clinical risk markers: bacterial STI history and recent HIV test (from the previous year). Incidence was further stratified by PrEP use in 2022/23.

The number of HIV-negative GBMSM attending SHS in England increased by 34% from 111,977 in 2014/15 to 149,904 in 2022/23, of whom repeat testers were 34% (37,576) in 2014/15 and 38% (56,896) in 2022/23. HIV incidence reduced by 93% overall (1.77/100py [95 CI 1.61-1.94] in 2014/15 to 0.12/100py [0.09-0.16] in 2022/23). Incidence reduced by at least 89% in all groups irrespective of clinical risk markers: with bacterial STI history (3.68/100py [3.17-4.27] to 0.26 [0.18-0.38]); the subset with a rectal bacterial infection (5.18/100py [4.13-6.49] to 0.58 [0.36-0.93]), and with a recent HIV test (1.93/100py [1.66-2.24] to 0.08 [0.05-0.13]). In 2022/23, using PrEP reduced HIV incidence by 86% (using PrEP: 0.05/100py [0.03-0.08]) vs not using PrEP: 0.36/100py [0.26-0.50]).

There was a sustained and large decline in HIV incidence among GBMSM, while incidence remains highest among those with a recent bacterial STI history. This analysis further highlights the real-world impact of PrEP and highlights the importance of equitable provision of HIV combination prevention interventions.

## Objectives

The English government committed to ending HIV transmission by 2030^1^, building on the observed large decline in new HIV diagnoses among gay, bisexual and other men who have sex with men (GBMSM) in England since 2015, with concurrent scaling up of HIV combination prevention interventionns^2^. However, measuring progress is not straightforward because underlying HIV transmission, migration and access to HIV testing, influence diagnosis trends, as people could be living with undiagnosed HIV for many years, not reflecting true incidence. While a decreasing trend in modelled HIV incidence in GBMSM overall has been reported^2,3^, less is known about HIV incidence trends among those with clinical risk markers^4-6^. Previous analyses used national surveillance data to derive HIV incidence in GBMSM attending sexual health services (SHS) based on previous HIV test and bacterial STI history^5,6^. We have replicated this method to derive annual HIV incidence between 2014/15 and 2022/23.

## Methods

### Data source

We used data from GUMCAD, the national STI surveillance system in England, that collects pseudonymised and depersonalised data of all SHS attendances with clinical and demographic information.

### Study population

GBMSM were defined as men who self-identified as gay or bisexual at least once over the study period. HIV-negative individuals or those not known to be living with HIV, (hereafter ‘HIV-negative’) were defined using clinical coding. All HIV-negative GBMSM aged 15 years and older attending a SHS between 2014/15 and 2022/23, are included in the analysis. Clinical markers of HIV risk included history of:

[a] bacterial STI diagnoses (chlamydia, gonorrhoea or infectious syphilis) in the previous year^4^,
[b] rectal bacterial STI diagnoses of chlamydia and gonorrhoea in the previous year (subset of [a])^4^, and
[c] recent HIV test in the previous year^7^ including [a] and [b] within this group.

To enable a comparison with previously published estimates of incidence^4,5^, an analysis using a wider range, including less commonly diagnosed bacterial STIs (donovanosis, chancroid, and lymphogranuloma venereum) and non-specific genital infection were included as a sensitivity analysis.

Only one HIV test is counted within a 42-day period, to avoid duplicate counts. The first attendance each year was used as the baseline date to determine these clinical risk markers.

### Estimating HIV incidence /Data analysis

Kaplan-Meier analysis was used to derive annual HIV incidence rates per 100 person-years (py) among GBMSM from 2014/15 to 2022/23, by clinical risk markers. We further stratified these groups by PrEP status in 2022/23 after the introduction of routinely commissioned PrEP in 2021. Individuals were defined as taking PrEP for up to 6 months since their last attendance where PrEP was prescribed if they were not reported to have stopped using PrEP.

We derived incidence among individuals with at least two HIV tests within 365 days (‘repeat testers’). Individuals were followed up for 365 days from their first HIV test each year until their last attendance (if they remain HIV-negative) or until they were newly diagnosed with HIV. The first HIV test each year was to confirm their HIV-negative status, and the subsequent HIV test to assess any change in HIV status during the follow-up period. Those who maintained their HIV-negative status and continued to be repeat testers in the following year were included in the cohort of the following year.

## Results

The number of HIV-negative GBMSM attending SHS in England increased by 34% from 111,977 in 2014/15 to 149,904 in 2022/23. Of 111,977 attendees in 2014/15, 16% (17,950) were diagnosed with bacterial STI, 5% (5,553) diagnosed with rectal bacterial STI, and 23% (25,341) had a recent HIV test in the previous year. Of 149,904 attendees in 2022/23, 20% (29,762) were diagnosed with bacterial STI, 6% (9,168) with rectal bacterial STI, and 26% (38,537) with a recent HIV test in the previous year (supplementary table 1a). Of HIV-negative GBMSM diagnosed with bacterial STIs at SHS in England, the majority were diagnosed with chlamydia, gonorrhoea or infectious syphilis: 80% (17,950/22,364) in 2014/15 and 93% (29,762/32,168) in 2022/23.

In 2022/23, 38% (56,896/149,904) of attendees were repeat testers, contributing 40,394py, and 49 were newly diagnosed with HIV with an overall incidence of 0.12/100py (95%CI 0.09-0.16); this was a 93% reduction from 1.77/100py (1.61-1.94) in 2014/15. By clinical risk markers, HIV incidence rates in 2022/23 was highest among group [b]: those diagnosed with rectal bacterial STIs in the past year (subset of [a]) (0.58/100py [0.36-0.93]). Compared to HIV incidence in 2014/15, this was a reduction by 89% from (5.18/100py [4.13-6.49]) in those diagnosed with rectal bacterial STI (Figure 1).

**Figure 1:**
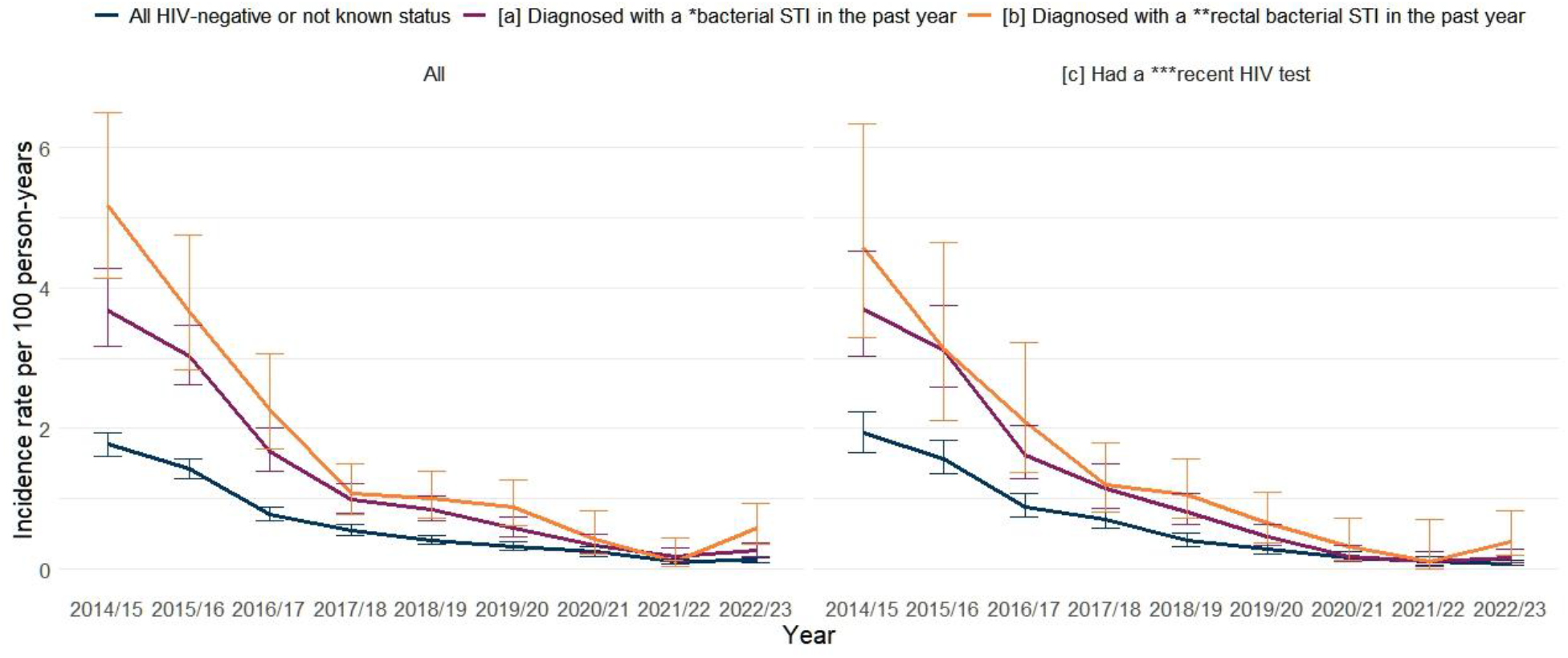
Annual HIV incidence rates among GBMSM attending sexual health services (SHS) in England, 2014/15 - 2022/23.

In 2022/23, after stratifying by PrEP use, HIV incidence rates was 0.36/100py (0.26-0.50) among non-PrEP users and 0.05/100py (0.03-0.08) among PrEP users (86% risk reduction). By other clinical risk markers, GBMSM taking PrEP had a greater reduction in incidence in groups [a]: with bacterial STI history and [b]: subset with rectal bacterial STI history (figure 1, supplementary table 1d).

## Conclusion

There was a steep and sustained decrease in HIV incidence by 93% in GBMSM between 2014/15 and 2022/23, with the most recent incidence at 0.12/100py. HIV incidence remained higher among GBMSM with a history of bacterial STIs, which further supports its reliability as a marker for increased HIV risk. These HIV incidence trends broadly align with the trends seen in the observed number of new HIV diagnoses in GBMSM, and modelled estimates of HIV incidence using a CD4 back-calculation method^2,3^. While most (up to 60%) of new HIV diagnoses in GBMSM are made in SHS^8^, these HIV incidence estimates derived using GUMCAD data are restricted to GBMSM attending SHS, and are therefore likely to over-estimate HIV incidence in all GBMSM.

Our analysis demonstrated further evidence of the effectiveness of PrEP for HIV prevention among GBMSM with an 86% reduction in HIV incidence among PrEP users, which matches the 86% reduction observed in the PROUD trial^9^ and the 86.8% reduction in the PrEP Impact trial^10^ in England. The PROUD trial was launched in 2012 and enrolled over 500 participants by 2014^9^. This was soon followed by the Impact trial in 2017, a larger scale implementation trial with over 20,000 GBMSM participants by 2020^10^. The routine commissioning of PrEP provision nationwide to those at risk of HIV, commenced in late 2020. By 2023, 83,210 GBMSM were reported to be using PrEP in England^3^. The impact of PrEP on HIV incidence over time likely commenced and continued through both the PROUD trial and the PrEP Impact trial because these trials included GBMSM at risk of HIV.

Frequent HIV testing, in part facilitated through the guidelines supporting the provision of PrEP, would also have helped reduce transmission by ensuring people who acquire HIV to be rapidly diagnosed and treated, to achieve viral suppression (an undetectable viral load), thereby minimising the risk of transmission. England achieved both the 2020 UNAIDS 90-90-90 target (i.e. 90% of those living with HIV to be diagnosed; 90% of those diagnosed to receive ART; 90% of those on ART to be virally suppressed) by 2016 and the 2030 95-95-95 target by 2020^1^. However, there is evidence of inequalities in the uptake of HIV combination prevention as, by ethnicity and region, the decline in new HIV diagnoses are comparatively lower among GBMSM ethnic minorities (excluding white minorities) living outside London^3^.

This analysis uses a comprehensive, large national surveillance dataset and identical statistical methods to previously published HIV incidence measures up to 2014^4-6^.

However, the depersonalised nature of the dataset prevents linking people who access healthcare from multiple SHS which may underestimate incidence, while defining PrEP status at censorship instead of at the start of follow-up may overestimate incidence.

Additionally, some of the reduction in incidence between 2018 and 2021 would reflect the fact that 20,000 GBMSM were taking PrEP through the PrEP Impact trial over that period.

While the CD4 back calculation method^3^ also shows decreasing trends in incidence since 2015, the approximate reduction in incidence is halved^3^ (0.33/100py-0.17/100py) compared to 93% (1.77/100py-0.12/100py) in this analysis. This reflects a difference in the settings of diagnosis data. The CD4 model includes all men diagnosed in any setting while our cohort is restricted to GBMSM accessing SHS and is therefore biased to men of higher HIV risk. The CD4 model also allows adjustment for probability of diagnosis; in our analysis we did not adjust for differences in HIV testing frequency between individuals; GBMSM at increased risk of HIV are likely to test more frequently and be diagnosed earlier, overestimating incidence. Conversely, those who test less frequently may have the infection undetected for longer periods, leading to an underestimation of incidence using the CD4 back-calculation method.

The decline in HIV incidence between 2014/15 and 2022/23 across clinical risk subgroups of GBMSM highlights the success of combination prevention including access to PrEP for this population. However, trends in HIV diagnoses suggest widening inequalities in reducing transmission within GBMSM. To maintain this progress and reach zero HIV transmission in England by 2030, it is important that HIV combination prevention interventions are equitably provided to all populations at increased risk of HIV.

## Data Availability

The data that support the findings of this analysis have been assessed by the UK Health Security Agency Office for Data Acquisition and Release as having sensitive personal information and are therefore not publicly available to protect participant privacy. However, some aggregate data may be available upon reasonable request from the UKHSA. Requests can be directed to.

