## Supplemental Files for "Annual HIV incidence among gay, bisexual and other men who have sex with men in England, 2014 to 2023: A prospective cohort analysis using national surveillance data"

### Supplementary tables/figures

#### Table 1a-1d: Number, proportion and HIV incidence rates of GBMSM attending SHS, by clinical risk markers.

| **Table 1a. Number and proportion of GBMSM attending SHS, by clinical risk markers, in 2014/15, and in 2022/23** | | | | |
|---|---|---|---|---|
| **Clinical risk markers** | **2014/15 (n)** | **%** | **2022/23 (n)** | **%** |
| HIV negative or unknown (Total) | 111,977 | | 149,904 | |
| [a] Bacterial STI diagnoses in the previous year | 17,950 | 16% | 29,762 | 20% |
| [b] Recent rectal bacterial STI in the previous year (subset of [a]) | 5,553 | 5% | 9,168 | 6% |
| [c] history of HIV test in the previous 42-365 days including [a] and [b] within this group | 25,341 | 23% | 38,537 | 26% |
| Subset of [c] with bacterial STI diagnoses in the previous year | 6,712 | 6% | 13,554 | 9% |
| Subset of [c] with rectal bacterial STI in the previous year | 2,147 | 2% | 3,563 | 2% |
| **Table 1b. Number and proportion of GBMSM attending SHS, who were repeat testers, by clinical risk markers , in 2014/15, and in 2022/23** | | | | |
| **Clinical risk markers** | **2014/15 (n)** | **%** | **2022/23 (n)** | **%** |
| HIV negative or unknown (Total) | 37,576 | 34% | 56,896 | 38% |
| [a] Bacterial STI diagnoses in the previous year | 7,181 | 40% | 14,499 | 49% |
| [b] Recent rectal bacterial STI in the previous year (subset of [a]) | 2,255 | 41% | 4,035 | 44% |
| [c] history of HIV test in the previous 42-365 days including [a] and [b] within this group | 12,921 | 51% | 24,123 | 63% |
| Subset of [c] with bacterial STI diagnoses in the previous year | 3,756 | 56% | 9,200 | 68% |
| Subset of [c] with rectal bacterial STI in the previous year | 1,172 | 55% | 2,340 | 66% |
| **Table 1c. HIV incidence rates among GBMSM attending SHS, who were repeat testers, by clinical risk markers, in 2014/15, and in 2022/23** | | | | |
| **Clinical risk markers** | **2014/15** | | **2022/23** | |
| | **Repeat testers** | **Rates per 100 py (95%CI)** | **Repeat testers** | **Rates per 100 py (95%CI)** |
| HIV negative or unknown (Total) | 37,576 | 1.77 (1.61-1.94) | 56,896 | 0.12 (0.09-0.16) |
| [a] Bacterial STI diagnoses in the previous year | 7,181 | 3.31 (2.87-3.81) | 14,499 | 0.26 (0.18-0.38) |
| [b] Recent rectal bacterial STI in the previous year (subset of [a]) | 2,255 | 5.04 (4.02-6.32) | 4,035 | 0.58 (0.36-0.93) |

| | | | | |
|---|---|---|---|---|
| [c] history of HIV test in the previous 42-365 days including [a] and [b] within this group | 12,921 | 1.93 (1.66-2.24) | 24,123 | 0.08 (0.05-0.13) |
| Subset of [c] with bacterial STI diagnoses in the previous year | 3,756 | 3.40 (2.81-4.12) | 9,200 | 0.16 (0.09-0.29) |
| Subset of [c] with rectal bacterial STI in the previous year | 1,172 | 4.44 (3.21-6.16) | 2,340 | 0.39 (0.19-0.83) |

**Table 1d. HIV incidence rates among GBMSM attending SHS, who were repeat testers, by clinical risk markers, and PrEP use, in 2022/23**

| **Clinical risk markers** | **Repeat testers** | **Risk reduction** | **Not On PrEP** | **Using PrEP** |
|---|---|---|---|---|
| | | | **Rates per 100 py (95%CI)** | **Rates per 100 py (95%CI)** |
| HIV negative or unknown (Total) | 56,896 | 86% | 0.36 (0.26-0.50) | 0.05 (0.03-0.08) |
| [a] Bacterial STI diagnoses in the previous year | 14,499 | 92% | 0.99 (0.64-1.54) | 0.08 (0.04-0.17) |
| [b] Recent rectal bacterial STI in the previous year (subset of [a]) | 4,035 | 93% | 1.96 (1.16-3.31) | 0.13 (0.04-0.42) |
| [c] history of HIV test in the previous 42-365 days including [a] and [b] within this group | 24,123 | 87% | 0.31 (0.15-0.61) | 0.04 (0.02-0.09) |
| Subset of [c] with bacterial STI diagnoses in the previous year | 9,200 | 88% | 0.69 (0.31-1.55) | 0.08 (0.03-0.20) |
| Subset of [c] with rectal bacterial STI in the previous year | 2,340 | 83% | 1.26 (0.47-3.36) | 0.21 (0.07-0.64) |

#### Figure 1a-b: Diagram illustrating follow-up time, PrEP status and change in HIV status for HIV incidence calculation

##### 1a. Follow-up time (9 months) and PrEP status (PrEP user) of an individual who remained HIV-negative

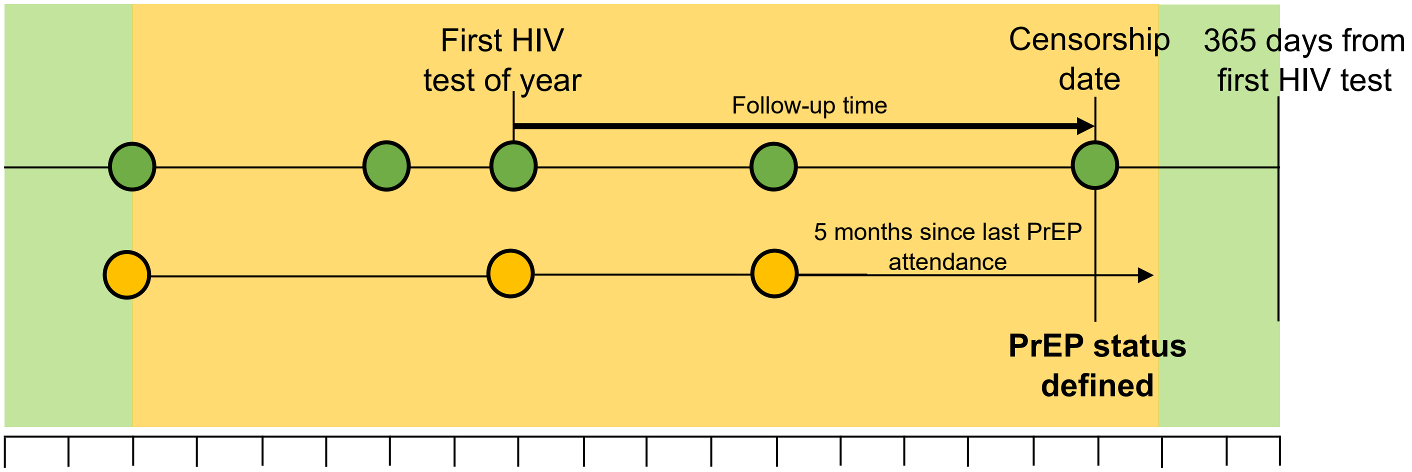


##### 1b. Follow-up time (8 months) and PrEP status (not a PrEP user) of an individual who was newly diagnosed with HIV

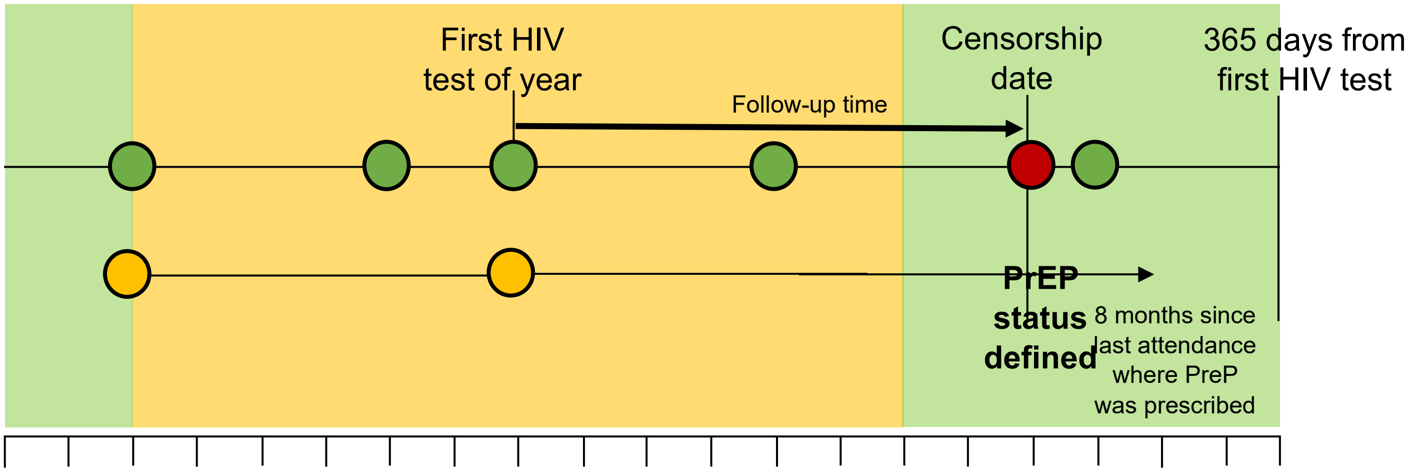


The follow-up period begins on a person's first HIV test of the year, and censorship occurs on the last attendance within 365 days of this test, or a new diagnosis of HIV.

Each person is defined as a PrEP user for 6 months from their most recent PrEP attendance, unless they have a PrEP stop code recorded in this time. Their PrEP status is defined by status at censorship

Diagram key

Period person is not on PrEP

Period person is on PrEP

Attendance

Attendance with PrEP started/continued code

Attendance, when diagnosed with HIV

#### Sensitivity analyses

##### Figure 2: Annual HIV incidence rates among GBMSM attending sexual health services (SHS) in England, 2014/15 - 2022/23 – sensitivity analyses which included a wider range of diagnosed bacterial STIs

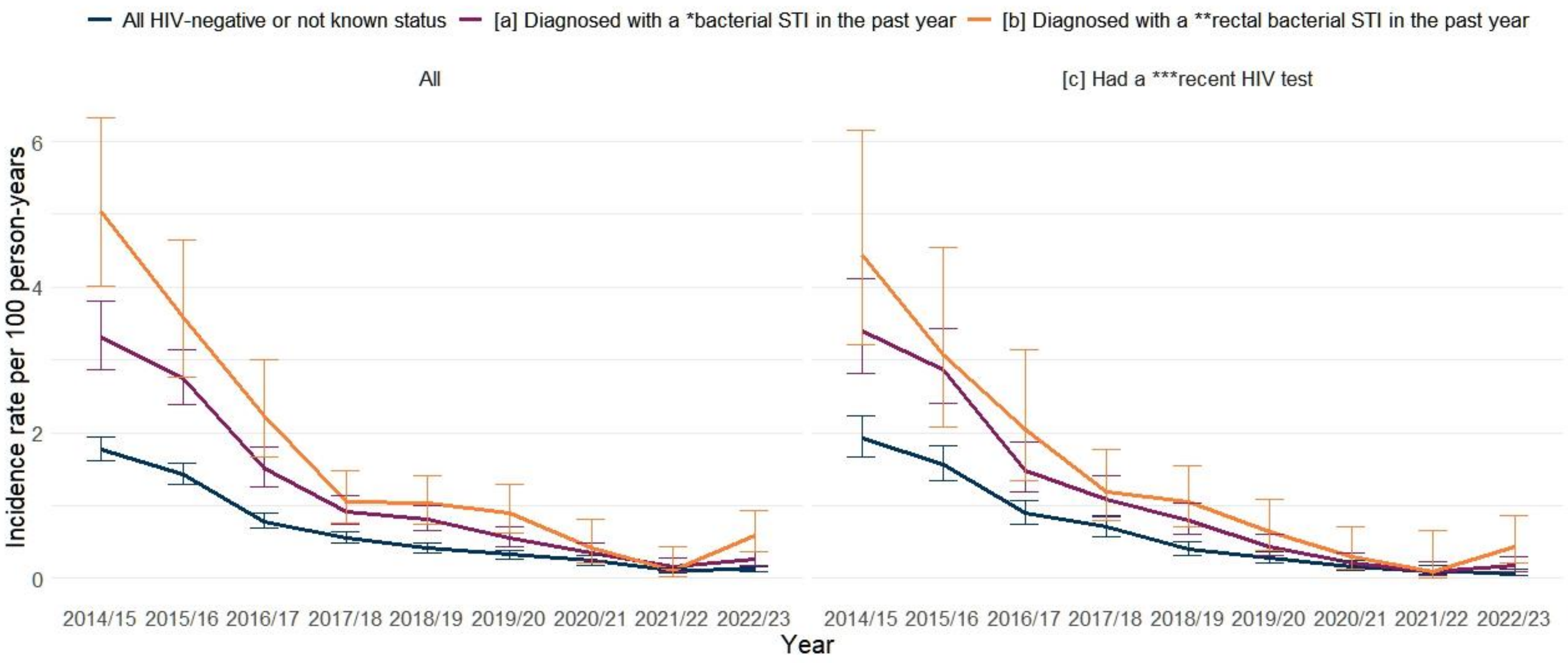


*Bacterial STIs included in this sensitivity analysis were diagnoses of chlamydia, gonorrhoea, infectious syphilis, non-specific genital infection (NSGI), donovanosis, chancroid, and Lymphogranuloma venereum (LGV).

**Rectal bacterial STIs included were for chlamydia, gonorrhoea and LGV diagnoses.

***Recent HIV test in the previous 42-365 days

#### Table 2a-2d: Number, proportion and HIV incidence rates of GBMSM attending SHS, by clinical risk markers, which includes both common and less commonly diagnosed bacterial STIs

| **Table 2a. Number and proportion of GBMSM attending SHS, by clinical risk markers, in 2014/15, and in 2022/23** | | | | |
|---|---|---|---|---|
| **Clinical risk markers** | **2014/15 (n)** | **2014/15 (%)** | **2022/23 (n)** | **2022/23 (%)** |
| HIV negative or unknown (Total) | 111,977 | | 149,904 | |
| [a] Bacterial STI diagnoses in the previous year | 22,364 | 20% | 32,168 | 21% |
| [b] Recent rectal bacterial STI in the previous year (subset of [a]) | 5,701 | 5% | 9,441 | 6% |
| [c] history of HIV test in the previous 42-365 days including [a] and [b] within this group | 25,341 | 23% | 38,537 | 26% |
| Subset of [c] with bacterial STI diagnoses in the previous year | 8,264 | 7% | 14,306 | 10% |
| Subset of [c] with rectal bacterial STI in the previous year | 2,199 | 2% | 3,693 | 2% |
| **Table 2b. Number and proportion of GBMSM attending SHS, who were repeat testers, by clinical risk markers, in 2014/15, and in 2022/23** | | | | |
| **Clinical risk markers** | **2014/15 (n)** | **2014/15 (%)** | **2022/23 (n)** | **2022/23 (%)** |
| HIV negative or unknown (Total) | 37,576 | 34% | 56,900 | 38% |
| [a] Bacterial STI diagnoses in the previous year | 8,704 | 39% | 15,326 | 48% |
| [b] Recent rectal bacterial STI in the previous year (subset of [a]) | 2,313 | 41% | 4,179 | 44% |
| [c] history of HIV test in the previous 42-365 days including [a] and [b] within this group | 12,921 | 51% | 24,123 | 63% |
| Subset of [c] with bacterial STI diagnoses in the previous year | 4,553 | 55% | 9,625 | 67% |
| Subset of [c] with rectal bacterial STI in the previous year | 1,203 | 55% | 2,430 | 66% |
| **Table 2c. HIV incidence rates among GBMSM attending SHS, who were repeat testers, by clinical risk markers, in 2014/15, and in 2022/23** | | | | |
| **Clinical risk markers** | **2014/15** | | **2022/23** | |
| | **Repeat testers** | **Rates per 100 py (95%CI)** | **Repeat testers** | **Rates per 100 py (95%CI)** |
| HIV negative or unknown (Total) | 37,576 | 1.77 (1.61-1.94) | 56,900 | 0.11 (0.08-0.15) |
| [a] Bacterial STI diagnoses in the previous year | 8,704 | 3.31 (2.87-3.81) | 15,326 | 0.16 (0.10-0.28) |
| [b] Recent rectal bacterial STI in the previous year (subset of [a]) | 2,313 | 5.04 (4.02-6.32) | 4,179 | 0.11 (0.03-0.43) |

| | | | | |
|---|---|---|---|---|
| [c] history of HIV test in the previous 42-365 days including [a] and [b] within this group | 12,921 | 1.93 (1.66-2.24) | 24,123 | 0.10 (0.06-0.17) |
| Subset of [c] with bacterial STI diagnoses in the previous year | 4,553 | 3.40 (2.81-4.12) | 9,625 | 0.10 (0.04-0.23) |
| Subset of [c] with rectal bacterial STI in the previous year | 1,203 | 4.44 (3.21-6.16) | 2,430 | 0.09 (0.01-0.65) |

**Table 2d. HIV incidence rates among GBMSM attending SHS, who were repeat testers, by clinical risk markers, and PrEP use, in 2022/23**

| **Clinical risk markers** | **Repeat testers** | **HIV risk reduction among PrEP users** | **Not On PrEP** | **Using PrEP** |
|---|---|---|---|---|
| | | | **Rates per 100 py (95%CI)** | **Rates per 100 py (95%CI)** |
| HIV negative or unknown (Total) | 56,900 | -86% | 0.36 (0.26-0.50) | 0.05 (0.03-0.08) |
| [a] Bacterial STI diagnoses in the previous year | 15,326 | -91% | 0.94 (0.62-1.45) | 0.08 (0.04-0.17) |
| [b] Recent rectal bacterial STI in the previous year (subset of [a]) | 4,179 | -91% | 1.90 (1.13-3.21) | 0.17 (0.06-0.46) |
| [c] history of HIV test in the previous 42-365 days including [a] and [b] within this group | 24,123 | -87% | 0.31 (0.15-0.61) | 0.04 (0.02-0.09) |
| Subset of [c] with bacterial STI diagnoses in the previous year | 9,625 | -89% | 0.75 (0.36-1.58) | 0.08 (0.03-0.19) |
| Subset of [c] with rectal bacterial STI in the previous year | 2,430 | -79% | 1.22 (0.46-3.25) | 0.26 (0.10-0.70) |
